# FACT- Frankfurt adjusted COVID-19 testing- a novel method enables high-throughput SARS-CoV-2 screening without loss of sensitivity

**DOI:** 10.1101/2020.04.28.20074187

**Authors:** Michael Schmidt, Sebastian Hoehl, Annemarie Berger, Heinz Zeichhardt, Kai Hourfar, Sandra Ciesek, Erhard Seifried

## Abstract

**Background:** In the pandemic, testing for SARS-CoV-2 by RT-PCR in one of the pillars on which countermeasures are based. Factors limiting the output of laboratories interfere with the effectiveness of public health measures. Conserving reagents by pooling samples in low-probability settings is proposed, but may cause dilution and loss of sensitivity.

**Methods:** We tested an alternate approach (FACT) by simultaneously incubating multiple respiratory swabs in a single tube. This protocol was evaluated by serial incubation of a respiratory swab in up to 10 tubes. The analytics validity of this concept was demonstrated in a five-sample mini pool set-up. It was consequently applied in the testing of 50 symptomatic patients (five-sample pools) as well as 100 asymptomatic residents of a nursing home (ten-sample pools).

**Results:** Serial incubation of a respiratory swab in up to 10 tubes did not lead to a significant decline in viral concentration. The novel FACT-protocol did not cause a false negative result in a five-sample mini-pool setup, with non-significantly differing Ct values between single sample and mini-pool NAT. In two routine applications, all mini pools containing positive patient samples were correctly identified.

**Conclusions:** Our proposed FACT-protocol did not cause a significant loss in analytic or diagnostic sensitivity compared to single sample testing in multiple setups. It reduced the amount of reagents needed by up to 40%, and also reduced hands-on time. This method could enhance testing efficiency, especially in groups with a low pretest-probability, such as systemically relevant professional groups.

## Introduction

SARS-CoV-2 is the causative agent of the novel lung disease COVID-19. With more than 1.3 million cases and almost 80 thousand deaths recorded worldwide by April 8^th^ 2020^1^, cases are still rising sharply in many parts of the world. Nations throughout the world are attempting to slow down the surge in cases by putting extensive countermeasures in place.

Infection may remain asymptomatic or pass with only minor symptoms, making a clinical diagnosis impossible in many cases.^2–4^ High viral titers in the upper airways during the first week of symptoms^5,6^ and presymptomatic transmission^7^ likely contributes to the difficulty containing the pandemic. In the struggle against the pandemic, the WHO recently urged nations to „test, test, test“.^8^ Detection of SARS-CoV-2 by nucleic amplification technologies (NAT), such as PCR, in a nasopharyngeal or throat swab and/or lower respiratory specimen is the preferred method as recommended by the WHO.^9^

The unprecedented demand for NAT reagents and test kits has already led to shortages, obstructing the efforts to combat COVID-19. Another factor limiting the output of laboratories is the availability of qualified staff. Furthermore, especially in low-income settings, where the threat by COVID-19 is no less imminent, cases may go undetected when tests are too expensive.

To make testing for SARS-CoV-2 more efficient, sample pooling has been proposed, and recently applied in a retrospective analysis.^10^ Dilution effects leading to a loss in diagnostic sensitivity is a concern in this strategy, when sample solutions are pooled. Here, we evaluated an alternate FACT protocol (Frankfurt adjusted COVID-19 testing) that allows a ten times higher number of SARS-CoV-2 tests.

## Material and Methods

### Alternate sample-pool protocol

We applied a novel protocol to NAT testing of respiratory swabs for SARS-CoV-2: Respiratory swabs were first incubated in a reference tube containing 4.3 ml of guanidinium thiocyanate buffer solution for 5 minutes with constant agitation. Consequently, all swabs used for mini-pooling are removed and collectively placed in one new single media pool tube containing 2 ml of guanidinium thiocyanate buffer, the mini-pool tube, under constant agitation for 5 minutes (Figure 1). The swabs are then removed from the mini-pool tube, which proceeds to NAT testing. Reference tubes are stored at 2-8°C until mini pool NAT analysis is completed. In case of a negative result in the mini pool NAT, each sample in the mini-pool receives a negative result. If the result of a mini pool NAT is positive, individual SARS-CoV-2 NATs are carried out from the reference tubes.

**Figure 1:**
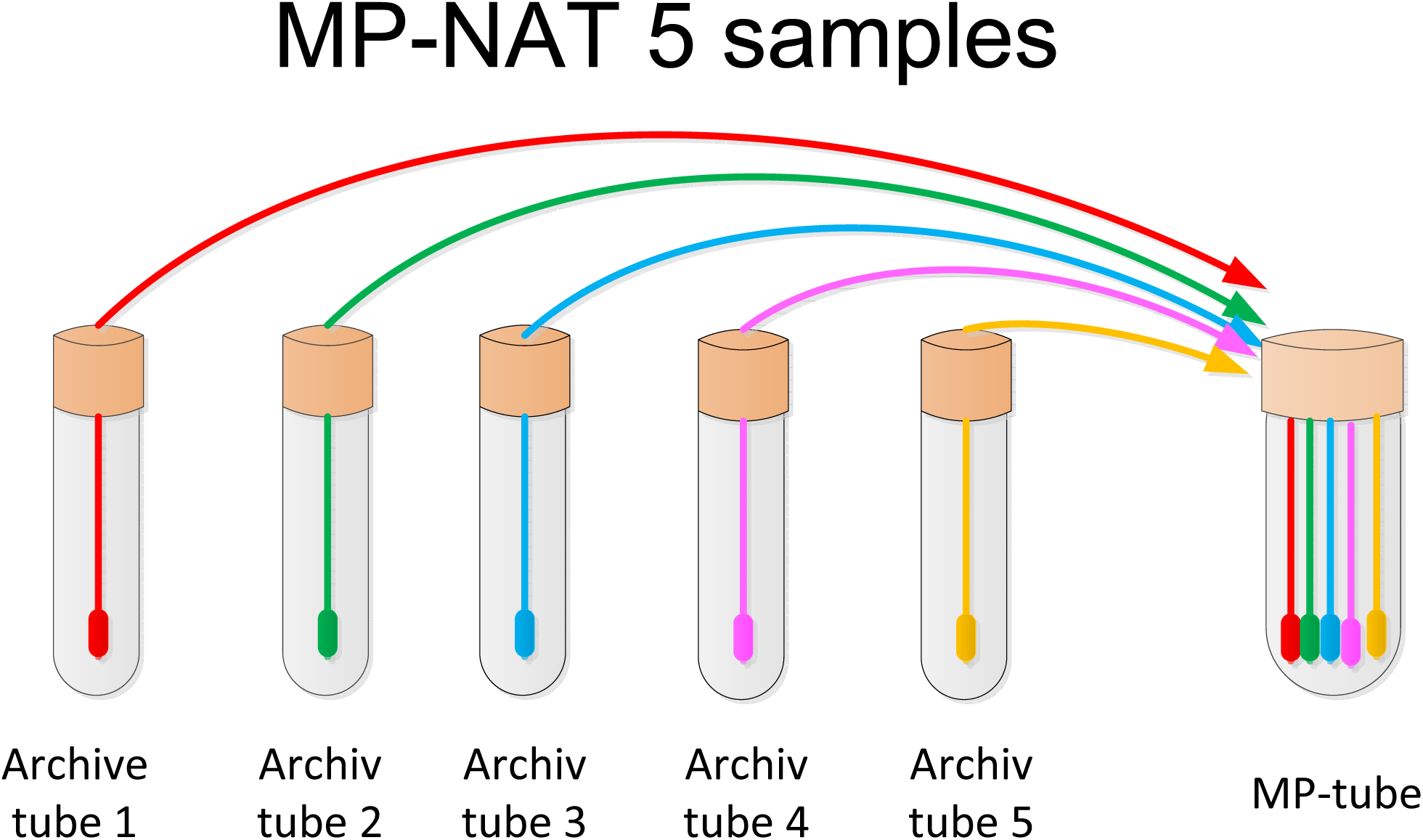
Mini-pool NAT method. Swaps were incubated first in a reference tube followed by five minute incubation in the mini-pool tube. SARS CoV-2 virus concentration did not differ significantly between both samples.

### NAT by RT-PCR

NAT was performed by Realtime-PCR (RT-PCR) on the Roche Cobas® 6800 or Roche Cobas® 8800 instrument. The sample input volume was 400µl. Amplification was done in a multiplex CE certified assay in the ORF region as well as in the E-gene. All samples were tested in accordance to the instruction for use from the manufacturer (Roche Diagnostics, Mannheim, Germany).

### Evaluation

The concept was assessed in three setups, and the diagnostic value was evaluated in practical application in symptomatic patients as well as in a screening procedure.

## Results

First, to evaluate for suitability of different mini-pool sizes, swabs were contaminated with a defined SARS-CoV-2 virus concentration of 1×10^4^ copies/ml, and then placed in a series of 10 tubes with lysis buffer for 5 minutes each. The virus concentration in each tube was determined. The results were examined for significant increase in Ct values in the succession of tubes, which would signify loss of sensitivity. We did not observe a significant difference in the semi-quantitative viral load between the first tube (representing individual sample testing) and the tenth tube. The largest observed difference in Ct value was 1.73 and 2.23 for ORF- and E-gene, respectively (table 1), which we consider not significant.

**Table 1:** Incubation of a SARS CoV-2 contaminated swap with 10^4^ copies/ml sequentially into ten sample tubes with lysis buffer

|  | Target 1 | Target 2 |
| --- | --- | --- |
| Tube 1 | 30.37 | 29.91 |
| Tube 2 | 30.59 | 30.29 |
| Tube 3 | 30.6 | 30.26 |
| Tube 4 | 31.32 | 30.95 |
| Tube 5 | 32.52 | 32.36 |
| Tube 6 | 32.19 | 32.01 |
| Tube 7 | 32.84 | 32.83 |
| Tube 8 | 32.61 | 32.68 |
| Tube 9 | 32.21 | 32.01 |
| Tube 10 | 32.1 | 32.14 |

The difference in Ct values between tube 1 and tube 10 was 1.73 and 2.23 for the ORF region and the E-gene, respectively.

Next, we evaluated a five-sample mini pool in a proof-of-concept setup. Samples from a round robin test provider (INSTAND, Düsseldorf, Germany) with predetermined concentrations of SARS-CoV-2 were used. Four of the samples were incubated in a solution containing SARS-CoV-2, one was incubated in solution not containing virus. Each of the swabs was transferred to a five-sample mini-pool, in accordance with the protocol described above. Respiratory swabs from SARS-CoV-2 negative volunteers were used to complete the pools. Results of single sample and mini-pool testing were compared. We determined that all mini-pools containing a SARS-CoV-2 positive sample were correctly identified in the mini-pool protocol, independent of the virus concentration in the original sample. The mini-pool containing no SARS-CoV-2 positive sample was also true negative. When comparing the Ct values in the single sample and the mini-pool (table 2), the largest observed gap between Ct values was 0.87 (sample 4, in the E gene as well as ORF-region), which we consider not significant.

**Table 2:** Comparative Ct value results in a proof-of-concept approach with round robin trial samples

|  | ID-NAT | MP-NAT |
| --- | --- | --- |
| Sample ID | ORF-region | ORF-region |
| 1 | 28.02 | 27.25 |
| 2 | 30.60 | 30.40 |
| 3 | 33.41 | 33.22 |
| 4 | 36.33 | 35.46 |
| 5 | negative | negative |
| Sample ID | E gene | E gene |
| 1 | 28.51 | 28,01 |
| 2 | 31.42 | 31-23 |
| 3 | 34-40 | 34-55 |
| 4 | 37-98 | 37-11 |
| 5 | negative | negative |

Ct values did not differ significantly between the individual sample and mini-pool tube.

To evaluate the test in patients with a moderate likeliness of SARS-CoV-2 infection, 50 samples routinely sent in for SARS-CoV-2 testing of patients with clinical symptoms were randomly assigned to ten five-sample mini-pools. Both the reference tube and the mini-pool tube underwent NAT testing. Each of the four pools containing a positive sample was correctly identified in the mini-pool protocol. Mini-pools containing no positive sample were also correctly identified to be negative in the five sample mini-pool. Table 3 shows the comparative presentation of the Ct values from both methods. P-value for individuals sample and mini pool NAT was 0.299 and 0.354 for the ORF region and E-gene, respectively, which we consider not statistically significant.

**Table 3:** Statistical analysis of the novel mini pool protocol applied in the testing of 50 symptomatic patients. Comparative results of the four five-sample mini pools containing a SARS-CoV-2 positive sample (mini pools 1, 4, 5 and 6) are shown

| Pool number | ORF region |  | E gene |  |
| --- | --- | --- | --- | --- |
|  | Individual sample NAT | Mini pool NAT | Individual sample NAT | Mini pool NAT |
| 1 | 29.07 | 29.21 | 30.07 | 30.61 |
| 4 | 33.85 | 40.00 | 36.28 | 36.34 |
| 5 | 21.24 | 19.03 | 21.72 | 19.65 |
| 6 | 26.12 | 26.23 | 26.65 | 27.11 |

P-value was evaluated with the T-Test. The p-value was 0.299 and 0.354 between the combined Ct values from the individual sample NAT and mini-pool NAT of the ORF region and the E-gene, respectively.

In a second real-life application, 100 samples from asymptomatic residents of a nursing home were randomly assigned to ten mini-pools containing ten samples each. All five pools containing a total of eight positive samples were correctly identified. All five mini-pools containing no positive sample were also true negative. Ct-values did not differ significantly between mini-pool and the single sample testing (p-value for the ORF region and E gene were 0.44 and 0.46, respectively) (table 4).

**Table 4:**
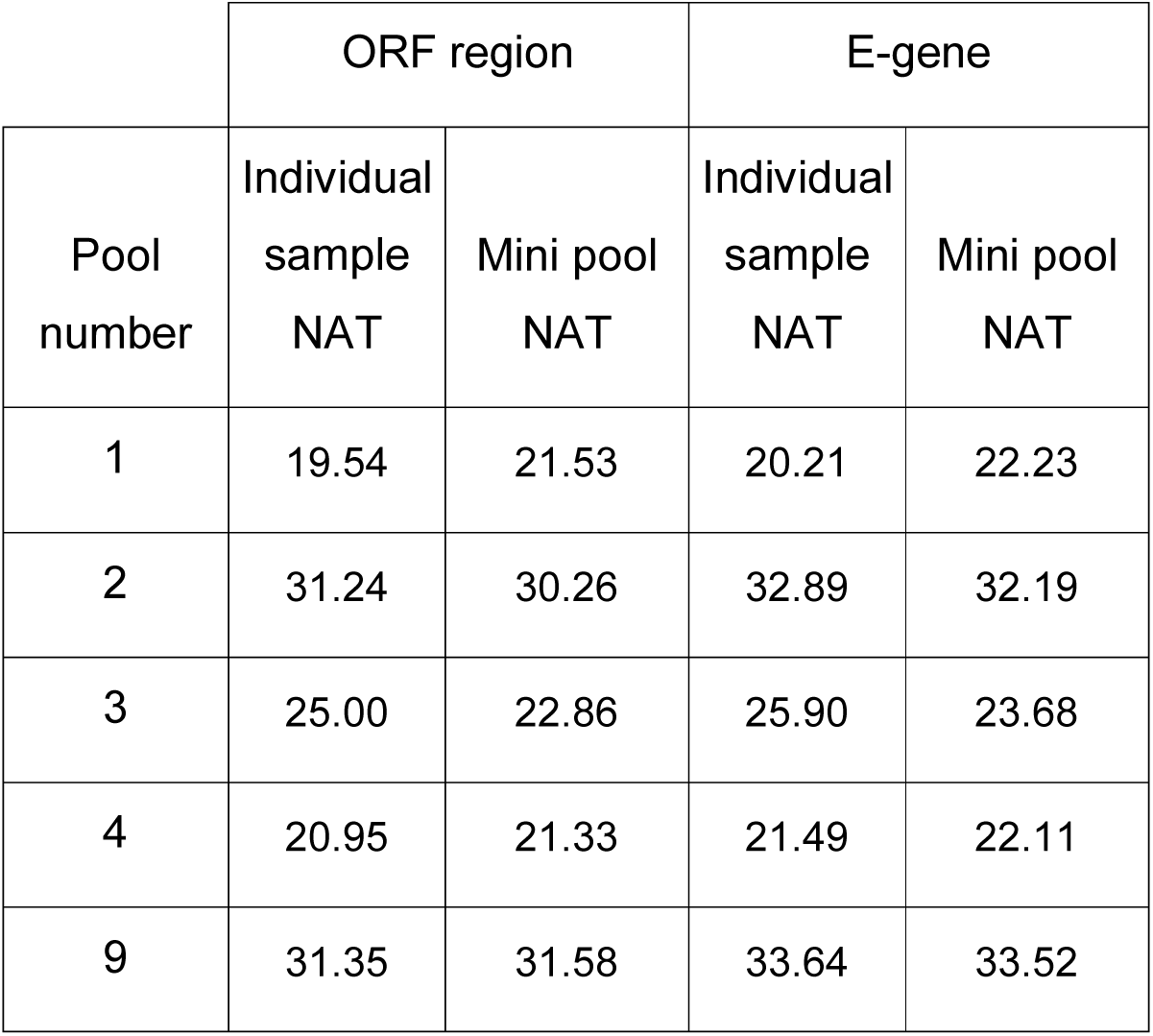
Statistical analysis of the mini-pool protocol applied in a nursing home screening. Comparative results of the five mini pools containing SARS-CoV-2 positive sample (mini pools 1, 2, 3, 4 and 9) are shown.

## Discussion

Increased test efficiency is eagerly awaited for SARS-CoV-2, as effective strategies to slow down the pandemic depend on early detection of cases, while a finite supply of reagents, qualified personnel and high costs interfere. To preserve reagents, reduce hands-on time and expenses, sample pooling is being proposed for settings with a low pre-test probability.^10,11^ This is usually conducted by pooling solutions containing the material that is to be tested. However, loss of sensitivity is a concern in this approach, as dilution occurs.

Here, we present an alternate protocol (FACT) that is based on incubation of a respiratory swab first in a single sample tube, and then again in a mini-pool tube. We detected no significant difference in the amount of virus detectable by NAT in the single sample and mini-pool tube. Therefore, by applying this protocol in the diagnostic process, no loss of diagnostic or analytical sensitivity would be expected, dismissing a main concern that might hinder implementation. We presume that our mini pooling process can be implemented for all NAT methods and all dry swaps. We applied the protocol in two routine scenarios, where the novel protocol was able to reduce to total number of required NAT tests by up to 40%, without loss of diagnostic sensitivity.

By putting this method into practice in the current SARS-CoV-2 pandemic, the number of samples that can be tested with a given amount of NAT reagents could immediately be increased in a sub-cohort with a low pretest probability, when it is not likely that pools must be resolved and samples tested individually, which would void the initial benefit. This could be especially useful when screening professional groups that are exposed to the virus while also posing a risk of spreading it, such as health care workers and emergency responders, or groups at risk, such as the elderly. This approach would not be efficient in a setting with high pre-test probability, where it would be likely that the individual samples would have to be retested. Here, a single sample test, or a smaller pool size would be advisable.

Mathematic models have recently also addressed the beneficial effects of conventional pooled testing strategies. A pre-print paper by Hanel et al. evaluated the optimal pool size for varying infection level in the population^11^, and suggested an optimal pool size of eleven for an infection level of 1%, which could lead to a 4 fold gain in efficiency. However, with rising infection levels, this gain also decreases. Further research on the optimal test size in varying population frequencies of infection is needed using the novel protocol.

Parallels can be drawn to transfusion medicine, where pooling blood samples is an established method to reduce transmission of viral infections, such as HIV-1, HIV-2, HAV, HBV, HCV, HEV, WNV, CMV and B19 these viruses^12^, among other areas of application.

In summary, an efficient mini-pool strategy is urgently required. The FACT-protocol presented in this paper can be implemented immediately worldwide and thus could represent an essential component in the fight against the SARS CoV-2 pandemic.

## Data Availability

All data are available reffered to in the mansucript

## References

1 The World Health Organization (WHO). Coronavirus disease (COVID-19) outbreak situation. https://www.who.int/emergencies/diseases/novel-coronavirus-2019 (accessed March 31st, 2020).

2 Rothe C, Schunk M, Sothmann P, et al. Transmission of 2019-nCoV Infection from an Asymptomatic Contact in Germany. N Engl J Med 2020; 382: 970–71. https://doi.org/10.1056/NEJMc2001468.

3 Hoehl S, Rabenau H, Berger A, et al. Evidence of SARS-CoV-2 Infection in Returning Travelers from Wuhan, China. N Engl J Med 2020; 382: 1278–80. https://doi.org/10.1056/NEJMc2001899.

4 Song J-Y, Yun J-G, Noh J-Y, Cheong H-J, Kim W-J. Covid-19 in South Korea - Challenges of Subclinical Manifestations. N Engl J Med 2020. https://doi.org/10.1056/NEJMc2001801.

5 Wölfel R, Corman VM, Guggemos W, et al. Virological assessment of hospitalized patients with COVID-2019. Nature 2020. https://doi.org/10.1038/s41586-020-2196-x.

6 Zou L, Ruan F, Huang M, et al. SARS-CoV-2 Viral Load in Upper Respiratory Specimens of Infected Patients. N Engl J Med 2020; 382: 1177–79. https://doi.org/10.1056/NEJMc2001737.

7 Wei WE, Li Z, Chiew CJ, Yong SE, Toh MP, Lee VJ. Presymptomatic Transmission of SARS-CoV-2 — Singapore, January 23–March 16, 2020. MMWR Morb. Mortal. Wkly. Rep. 2020; 69. https://doi.org/10.15585/mmwr.mm6914e1.

8 The World Health Organization (WHO). WHO Director-General’s opening remarks at the media briefing on COVID-19 - 16 March 2020. https://www.who.int/dg/speeches/detail/who-director-general-s-opening-remarks-at-the-media-briefing-on-covid-19---16-march-2020 (accessed March 31st, 2020).

9 The World Health Organization (WHO). Laboratory testing for coronavirus disease (COVID-19) in suspected human cases. Interim guidance 19 March 2020 (accessed April 9th, 2020).

10 Hogan CA, Sahoo MK, Pinsky BA. Sample Pooling as a Strategy to Detect Community Transmission of SARS-CoV-2. JAMA 2020. https://doi.org/10.1001/jama.2020.5445.

11 Hanel R, Thurner S. Boosting test-efficiency by pooled testing strategies for SARS-CoV-2, 2020.

12 Roth WK, Weber M, Seifried E. Feasibility and efficacy of routine PCR screening of blood donations for hepatitis C virus, hepatitis B virus, and HIV-1 in a blood-bank setting. The Lancet 1999; 353: 359–63. https://doi.org/10.1016/S0140-6736(98)06318-1.

